# Severe outcomes in unvaccinated COVID-19 cases <18 years during different variant waves in Norway

**DOI:** 10.1101/2022.03.29.22273093

**Authors:** Robert Whittaker, Margrethe Greve-Isdahl, Håkon Bøås, Pål Suren, Eirik Alnes Buanes, Lamprini Veneti

## Abstract

**Objectives:** We used linked individual-level data from national registries to compare the risk of severe outcomes among unvaccinated COVID-19 cases <18 years between waves of the SARS-CoV-2 Alpha, Delta and Omicron variants in Norway.

**Methods:** Our outcomes were hospitalisation with acute COVID-19 or multisystem inflammatory syndrome in children (MIS-C). We calculated adjusted risk ratios (aRR) with 95% confidence intervals (CIs) using multivariable log-binomial regression, adjusting for variant wave, demographic characteristics and underlying comorbidities.

**Results:** We included 10,538 Alpha (21 hospitalised with acute COVID-19, 7 MIS-C), 42,362 Delta (28 acute COVID-19, 14 MIS-C) and 82,907 Omicron wave cases (48 acute COVID-19, 7 MIS-C). The risk of hospitalisation with acute COVID-19 in cases <1 year was lower in the Delta (aRR: 0.28, 95% CI: 0.16–0.89) and Omicron wave (aRR: 0.41, 95% CI: 0.20–0.81), compared to the Alpha wave. We found no difference in the risk for this outcome for Omicron compared to Delta in any age group. The risk of MIS-C was lower in the Omicron wave compared to the Alpha (aRR: 0.09, 95% CI: 0.03–0.27) and Delta wave (aRR: 0.26, 95% CI: 0.10–0.63).

**Conclusions:** We found no evidence of a difference in the risk of hospitalisation due to acute COVID-19 among unvaccinated cases <18 years for Omicron compared to Delta, but a reduced risk among cases <1 year in Omicron and Delta waves, compared to Alpha. Results also suggest a decrease in the risk of MIS-C in the Omicron wave compared to the Alpha and Delta waves.

**Article Summary:** We compare the risk of severe outcomes in unvaccinated COVID-19 cases <18 years between waves of the SARS-CoV-2 Alpha, Delta and Omicron variant in Norway.

**What’s Known on This Subject:** Currently, limited evidence suggests no clear difference in the risk of severe disease outcomes among children infected with different SARS-CoV-2 variants. The risk of multisystem inflammatory syndrome in children following infection with the Omicron variant has not been quantified.

**What This Study Adds:** We find a lower risk of hospitalisation due to acute COVID-19 among cases <1 year in the Delta and Omicron waves compared to the Alpha wave, and a lower risk of multisystem inflammatory syndrome in the Omicron wave, in Norway.

## Introduction

From late 2020, the emergence and global spread of variants of concern (VOC) (1) of severe acute respiratory syndrome coronavirus 2 (SARS-CoV-2) have shaped the epidemiology of, and ongoing response to, the coronavirus disease (COVID-19) pandemic. More transmissible VOC have successively superseded their predecessors influencing transmission dynamics, viral virulence and vaccine effectiveness (2-8). The emergence of the Omicron variant (Phylogenetic Assignment of Named Global Outbreak Lineages (Pangolin) designation B.1.1.529) in November 2021 instigated a new wave of infections globally (9).

Most children and adolescents with COVID-19 experience an asymptomatic or mild disease course. However, a small proportion may develop severe disease that requires hospitalisation. This may be due to acute COVID-19 or multisystem inflammatory syndrome in children (MIS-C), a postinfectious complication of SARS-CoV-2 infection. Death is rare (10, 11).

There is currently limited evidence on whether the relative severity of COVID-19 in children differs when infected with different VOC. Previous studies from England (5) and Denmark (12) have not found clear evidence that the risk of hospitalisation among children and adolescents infected with Omicron differed compared to the Delta variant (Pangolin designation B.1.617.2). Preliminary data from the United States have suggested an increase in upper respiratory complications among children since Omicron became dominant (13, 14). Some studies comparing the risk of hospitalisation for the Delta variant to earlier VOC based on reported cases have presented descriptive data for younger age groups, without providing age-specific risk estimates (15, 16). Others have been restricted to hospital cohorts, with the reported findings inconsistent between settings (17-19). A study from Denmark reported that the risk of MIS-C did not change among unvaccinated cases during a wave of Delta infections compared to when the wild-type SARS-CoV-2 variant was circulating (20).

Since the beginning of 2021, Norway has experienced three waves of COVID-19 cases when different VOC were dominant; Alpha (pangolin designation B.1.1.7) (4), Delta (21) and Omicron (8). We used linked individual-level data from national registries to compare the risk of severe outcomes among unvaccinated COVID-19 cases <18 years between these three variant waves.

## Methods

### Study setting

Norway (population 5.4 million, of whom 1.1 million <18 years) has had a broad testing strategy for COVID-19 in children since autumn 2020. Four pillars of the national pandemic response have been testing, isolation, contact tracing and quarantine. Through this framework SARS-CoV-2 tests have been available free of charge for everyone, including those with mild or no symptoms, close contacts and individuals in quarantine. Routine bi-weekly screening of school children with rapid antigen tests in areas with high transmission was undertaken for secondary school students from late-August 2021 and for primary school students from November 2021 until January 2022. Positive rapid antigen tests were confirmed with PCR. COVID-19 vaccine recommendations for children and adolescents in Norway and data on vaccination coverage by age group over time are presented in supplement A, section 1. From August 2021 there has been a general recommendation that all pregnant women in the second or third trimester get vaccinated. For women with underlying risk factors, vaccination was recommended in the first trimester (22).

### Data sources and study design

We obtained data through the Norwegian national preparedness registry for COVID-19 (23). The preparedness registry contains individual-level data from different central health registries, national clinical registries, and other national administrative registries. It covers all residents in Norway, and includes data on all laboratory-confirmed cases of COVID-19 in Norway and all hospitalisations, intensive care admissions and deaths among cases. Further details on the individual registries and data included in this study are presented in supplement A, section 2.

We conducted a cohort study, including reported cases of COVID-19 aged <18 years who tested positive from 15 March 2021 up to 30 January 2022, were unvaccinated at date of positive test and had also not previously been diagnosed with COVID-19, and had a national identity number registered. We extracted data up to 29 March 2022, ensuring a minimum 57 days of follow-up since last positive test.

### Definition of variant waves

The national laboratory database houses data on SARS-CoV-2 test results from all Norwegian microbiology laboratories. In Norway, SARS-CoV-2 variants are identified based on whole genome sequencing, Sanger partial S-gene sequencing or PCR screening targeting specific single nucleotide polymorphisms, insertions or deletions. The laboratory testing for variants of SARS-CoV-2 in Norway has been described in further detail elsewhere (24). Using data from the national laboratory database we identified different variant waves based on date of positive test. The distribution of different variants among cases <18 years over time in Norway is presented in Figure 1, with the underlying data available in supplement B. We defined the Alpha dominant wave as week 11–20 (15 March–23 May) 2021, the Delta dominant wave as week 35–48 (30 August–5 December) 2021 and the Omicron dominant wave as week 2–4 (10–30 January) 2022. In the Omicron wave, sublineage BA.1 was predominant (25).

**Figure 1.**
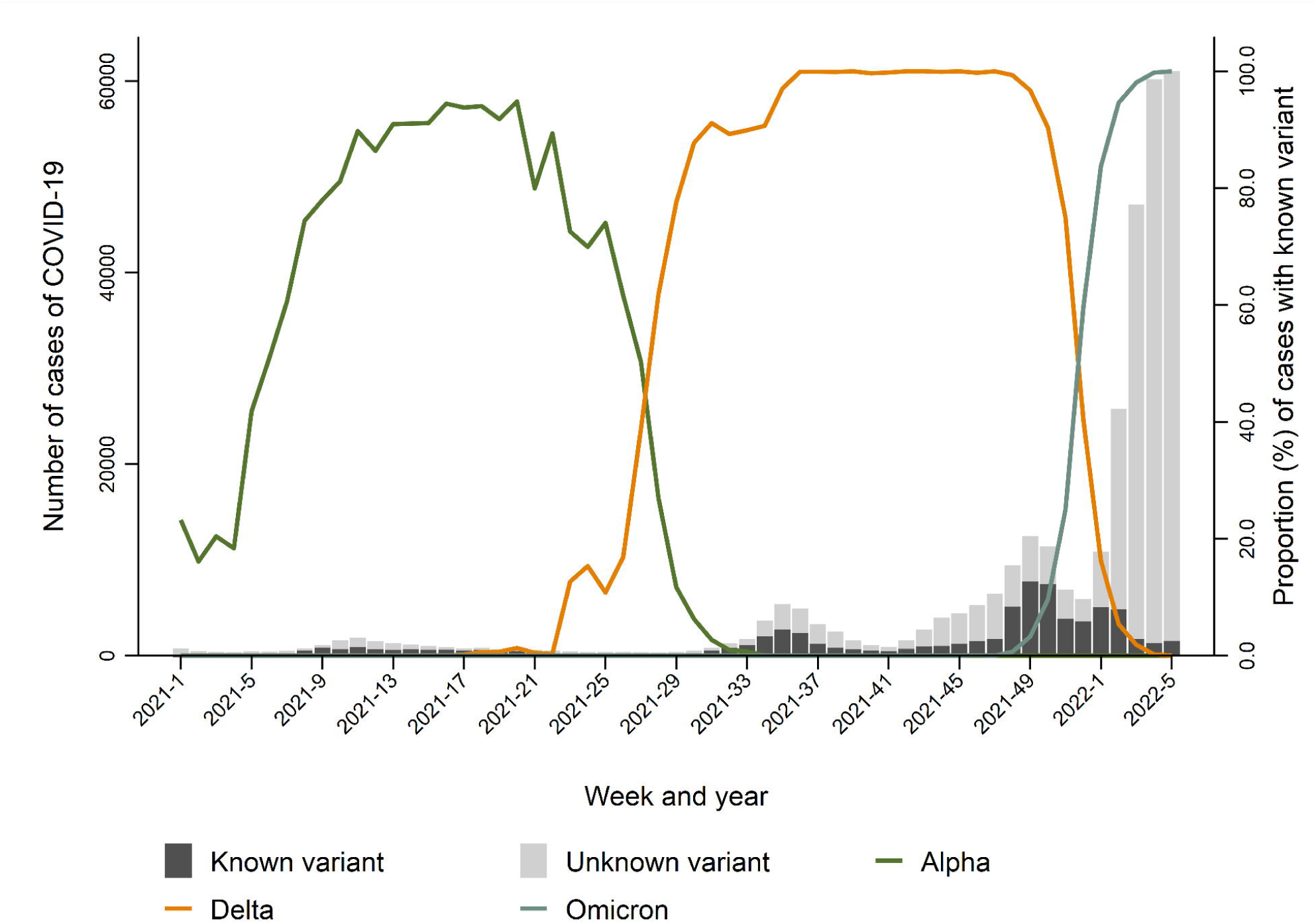
Reported COVID-19 cases aged <18 years in Norway with known and unknown variant (bars, left y-axis), and proportion of cases with known variant that were the Alpha, Delta or Omicron variant of concern (lines, right y-axis), by week, 1 January 2021 – 6 February 2022. The data behind the figure are available in supplement B. During the Alpha wave (week 11– 20 2021), the proportion of cases that were known to be infected with Alpha ranged from 86%–95% of cases with known variant, however, the vast majority of other cases with known variant were reported as ‘probable variant of concern’. In Norway, there was minimal circulation of other defined variants of concern (Beta, B.1.351 and Gamma, P.1), thus is it is reasonable to assume that most cases reported as ‘probable variant of concern’ were also the Alpha variant. Taking these cases into account the proportion infected with Alpha among those with known variant ranges from 94%–100% in the Alpha wave.

### Severity outcome definitions

Our severity outcomes of interest were: 1) admission to hospital with acute COVID-19 (regardless of main cause of admission) ≤14 days after positive test, 2) admission to hospital ≤14 days after positive test where acute COVID-19 was the reported main cause of admission and 3) admission to hospital with MIS-C, defined as patients registered with the International Classification of Diseases-10 diagnosis code U10.9. Clinical criteria for the diagnosis of MIS-C cases in Norway are based on the World Health Organisation case definition, as per national paediatric guidelines (26).

### Data analysis

We described cases by variant wave, severity outcome, demographic characteristics and underlying comorbidities. We also described other outcomes among hospitalised patients including length of stay (LoS) in hospital and admission to an intensive care unit (ICU), as well as all deaths in the study cohort. For our three severity outcomes, we calculated adjusted risk ratios (aRR) with 95% confidence intervals (CIs) using multivariable log-binomial regression. Explanatory variables to analyse differences in our outcomes included variant wave (Alpha, Delta or Omicron), age (as continuous or categorical variable), sex, country of birth, region of residence, and underlying comorbidities. The categorisation of explanatory variables is presented in Table 1 and further detailed in Supplement A, section 2. Explanatory variables were first checked in univariable models, and those with p<0.2 were further explored in multivariable models. Explanatory variables were further categorised in some models to best fit the data, for example a dichotomous variable for underlying comorbidities (yes/no). We maintained the variant wave variable in each multivariable analysis, even if not significant. We used Akaike Information Criteria and the likelihood ratio test to check model fit. We ran models for each variant combination (Delta vs. Alpha, Omicron vs. Alpha, Omicron vs. Delta) for all cases <18 years, as well as for the age subgroups <1, 1–11 and 12– 17 years. For the age group 12–17 years, we also conducted a sensitivity analysis, including vaccinated cases and reinfections, and adjusting for vaccination status (supplement A, section 3). Further, we validated the variant wave variable by running models for all COVID-19 cases reported in Norway, and comparing results to estimates based on cases with known variant (supplement A, section 4). Statistical analysis was performed in Stata version 16 (Stata Corporation, College Station, Texas, US).

**Table 1.**
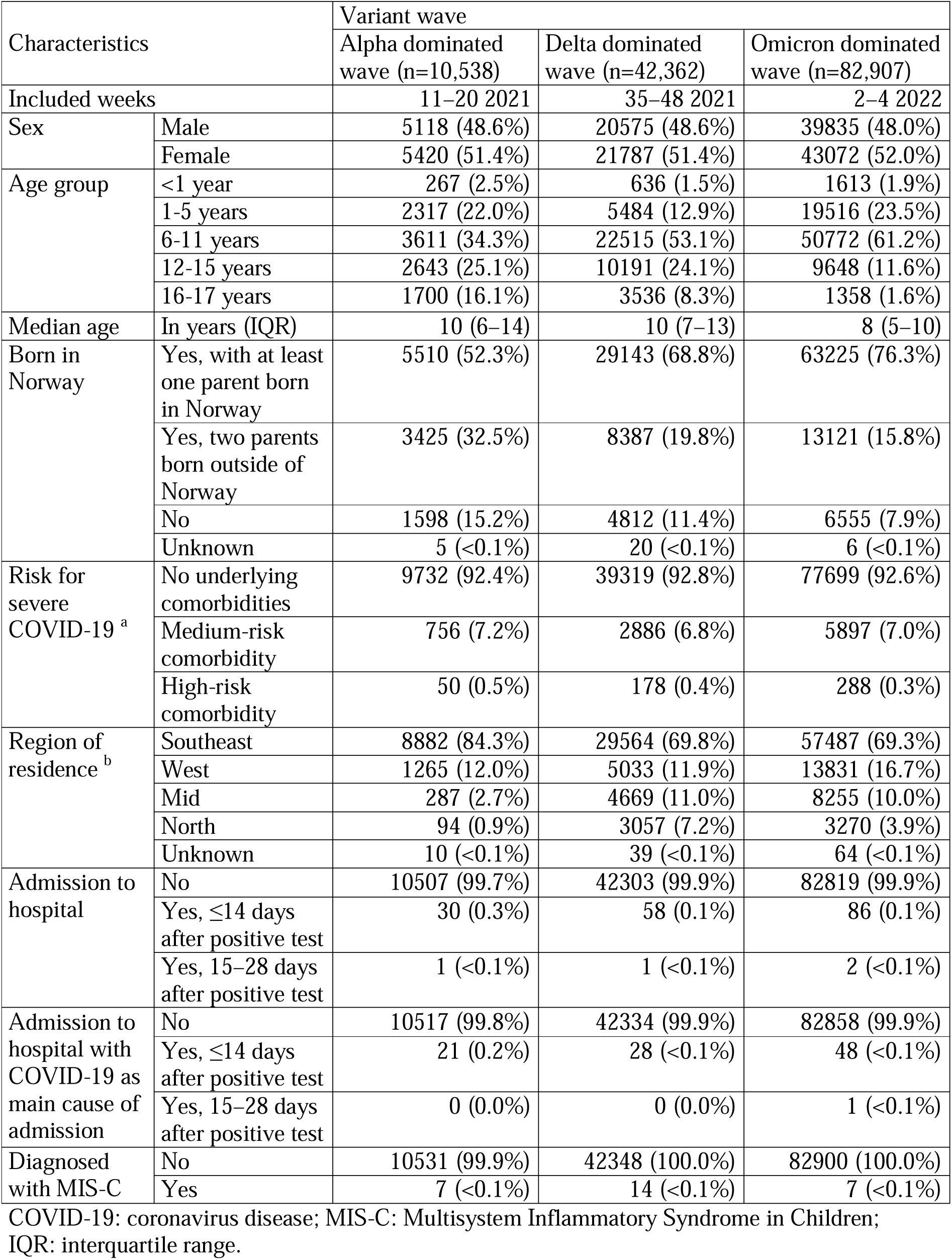
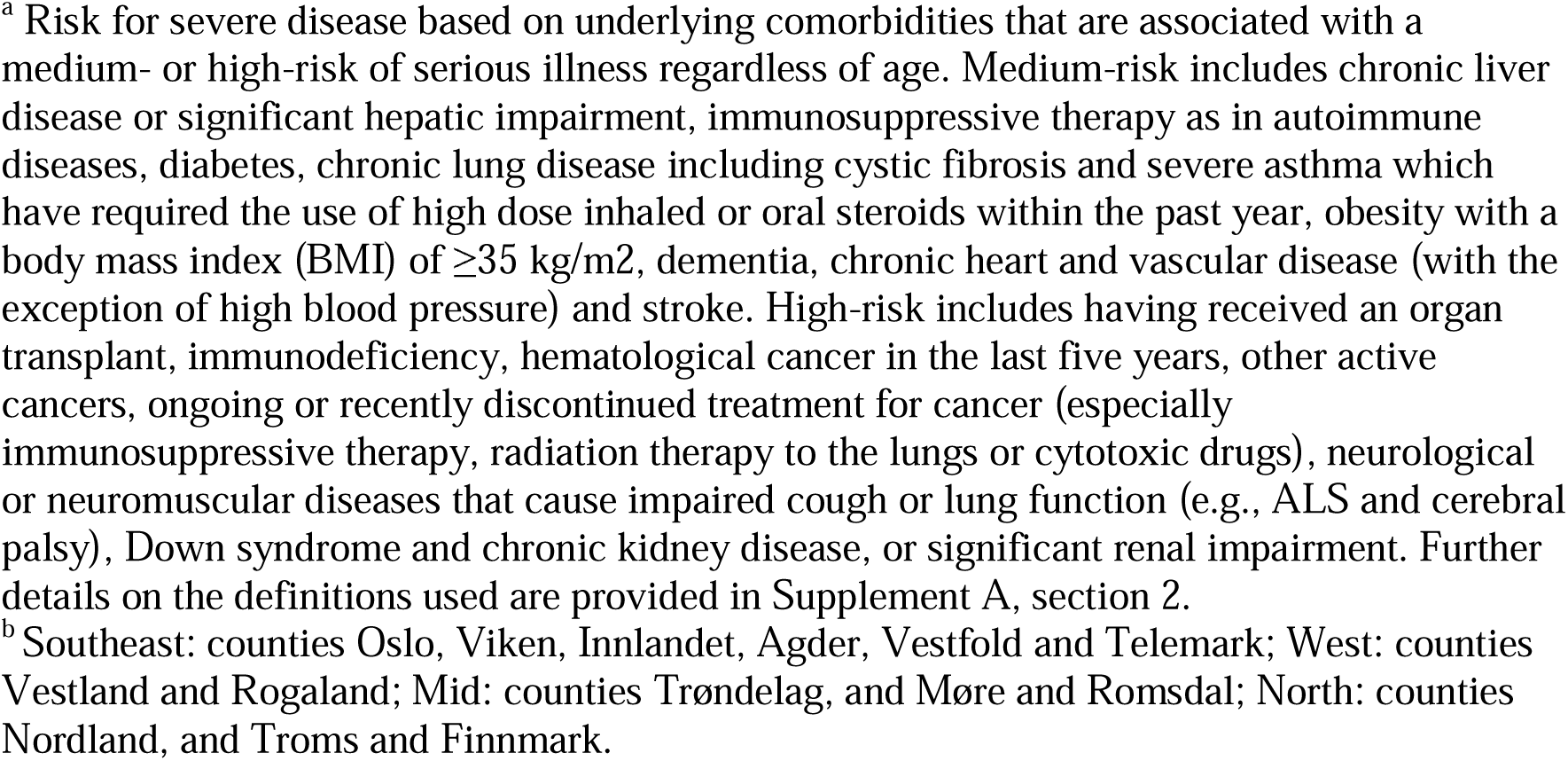
Characteristics of reported cases of COVID-19 aged <18 years who were unvaccinated and had also not previously been diagnosed with COVID-19, by variant wave, Norway, 15 March 2021 – 30 January 2022

| Characteristics |  | Variant wave |  |  |
| --- | --- | --- | --- | --- |
|  |  | Alpha dominated wave (n=10,538) | Delta dominated wave (n=42,362) | Omicron dominated wave (n=82,907) |
| Included weeks |  | 11–20 2021 | 35–48 2021 | 2–4 2022 |
| Sex | Male | 5118 (48.6%) | 20575 (48.6%) | 39835 (48.0%) |
|  | Female | 5420 (51.4%) | 21787 (51.4%) | 43072 (52.0%) |
| Age group | <1 year | 267 (2.5%) | 636 (1.5%) | 1613 (1.9%) |
|  | 1–5 years | 2317 (22.0%) | 5484 (12.9%) | 19516 (23.5%) |
|  | 6–11 years | 3611 (34.3%) | 22515 (53.1%) | 50772 (61.2%) |
|  | 12–15 years | 2643 (25.1%) | 10191 (24.1%) | 9648 (11.6%) |
|  | 16–17 years | 1700 (16.1%) | 3536 (8.3%) | 1358 (1.6%) |
| Median age | In years (IQR) | 10 (6–14) | 10 (7–13) | 8 (5–10) |
| Born in Norway | Yes, with at least one parent born in Norway | 5510 (52.3%) | 29143 (68.8%) | 63225 (76.3%) |
|  | Yes, two parents born outside of Norway | 3425 (32.5%) | 8387 (19.8%) | 13121 (15.8%) |
|  | No | 1598 (15.2%) | 4812 (11.4%) | 6555 (7.9%) |
|  | Unknown | 5 (<0.1%) | 20 (<0.1%) | 6 (<0.1%) |
| Risk for severe COVID-19 <sup>a</sup> | No underlying comorbidities | 9732 (92.4%) | 39319 (92.8%) | 77699 (92.6%) |
|  | Medium-risk comorbidity | 756 (7.2%) | 2886 (6.8%) | 5897 (7.0%) |
|  | High-risk comorbidity | 50 (0.5%) | 178 (0.4%) | 288 (0.3%) |
| Region of residence <sup>b</sup> | Southeast | 8882 (84.3%) | 29564 (69.8%) | 57487 (69.3%) |
|  | West | 1265 (12.0%) | 5033 (11.9%) | 13831 (16.7%) |
|  | Mid | 287 (2.7%) | 4669 (11.0%) | 8255 (10.0%) |
|  | North | 94 (0.9%) | 3057 (7.2%) | 3270 (3.9%) |
|  | Unknown | 10 (<0.1%) | 39 (<0.1%) | 64 (<0.1%) |
| Admission to hospital | No | 10507 (99.7%) | 42303 (99.9%) | 82819 (99.9%) |
|  | Yes, ≤14 days after positive test | 30 (0.3%) | 58 (0.1%) | 86 (0.1%) |
|  | Yes, 15–28 days after positive test | 1 (<0.1%) | 1 (<0.1%) | 2 (<0.1%) |
|  | Yes, ≥29 days after positive test | 0 (0.0%) | 0 (0.0%) | 0 (0.0%) |
| Admission to hospital with COVID-19 as main cause of admission | No | 10517 (99.8%) | 42334 (99.9%) | 82858 (99.9%) |
|  | Yes, ≤14 days after positive test | 21 (0.2%) | 28 (<0.1%) | 48 (<0.1%) |
|  | Yes, 15–28 days after positive test | 0 (0.0%) | 0 (0.0%) | 1 (<0.1%) |
|  | Yes, ≥29 days after positive test | 0 (0.0%) | 0 (0.0%) | 0 (0.0%) |
| Diagnosed with MIS-C | No | 10531 (99.9%) | 42348 (100.0%) | 82900 (100.0%) |
|  | Yes | 7 (<0.1%) | 14 (<0.1%) | 7 (<0.1%) |

COVID-19: coronavirus disease; MIS-C: Multisystem Inflammatory Syndrome in Children; IQR: interquartile range.
<sup>a</sup> Risk for severe disease based on underlying comorbidities that are associated with a medium- or high-risk of serious illness regardless of age. Medium-risk includes chronic liver disease or significant hepatic impairment, immunosuppressive therapy as in autoimmune diseases, diabetes, chronic lung disease including cystic fibrosis and severe asthma which have required the use of high dose inhaled or oral steroids within the past year, obesity with a body mass index (BMI) of $\geq 35$ kg/m<sup>2</sup>, dementia, chronic heart and vascular disease (with the exception of high blood pressure) and stroke. High-risk includes having received an organ transplant, immunodeficiency, hematological cancer in the last five years, other active cancers, ongoing or recently discontinued treatment for cancer (especially immunosuppressive therapy, radiation therapy to the lungs or cytotoxic drugs), neurological or neuromuscular diseases that cause impaired cough or lung function (e.g., ALS and cerebral palsy), Down syndrome and chronic kidney disease, or significant renal impairment. Further details on the definitions used are provided in Supplement A, section 2.
<sup>b</sup> Southeast: counties Oslo, Viken, Innlandet, Agder, Vestfold and Telemark; West: counties Vestland and Rogaland; Mid: counties Trøndelag, and Møre and Romsdal; North: counties Nordland, and Troms and Finnmark.

### Ethics

Ethical approval for studies on the risk of severe disease by SARS-CoV-2 variant based on data from the national preparedness registry for COVID-19 was granted by Regional Committees for Medical Research Ethics - South East Norway, reference number 249509.

## Results

### Study cohort

The total number of reported cases of COVID-19 aged <18 years was 10,620 during the Alpha wave, 53,724 during the Delta wave and 133,383 during the Omicron wave. Of these, 10,541 (99.3%) Alpha, 53,576 (99.7%) Delta and 133,042 (99.7%) Omicron cases had a known national identity number. Of these 10,538 (99.9%) Alpha, 42,362 (79.1%) Delta and 83,884 (62.3%) Omicron cases were included in the study cohort as they were unvaccinated at date of positive test and had not been previously diagnosed with COVID-19. Detailed characteristics of the study cohort by variant are presented in Table 1.

### Risk of hospitalisation

Overall, 174 (0.1%) cases were hospitalised ≤14 days after positive test, of which 97 with acute COVID-19 as main cause (0.07% of all cases in study cohort). Few additional admissions >14 days after positive test were observed (Table 1). Of the 97, 48 (50%) were <1 year of age, 44 (45%) were female and 83 (86%) had no registered comorbidity. Of the 48 <1 year, 32 (67%) were <3 months of age, and 42 (88%) were <6 months of age.

In the Alpha, Delta and Omicron waves, 21 (0.2%), 28 (0.07%) and 48 (0.06%) cases <18 years were hospitalised with acute COVID-19 as main cause ≤14 days after positive test, respectively (Table 1, Table 2). The risk of hospitalisation with acute COVID-19 as main cause was lower in the Delta (aRR: 0.53, 95% CI: 0.30–0.93) and Omicron wave (aRR: 0.40, 95% CI: 0.24–0.68) compared to the Alpha wave. This was explained by the age group <1 year (aRR Delta compared to Alpha: 0.28, 95% CI: 0.16–0.89; aRR Omicron compared to Alpha: 0.41, 95% CI: 0.20–0.81). Results also suggested a decreased risk in cases aged 1–11 years in the Omicron wave compared to the Alpha wave (aRR: 0.32, 95% CI: 0.13–0.83). We did not observe any difference in the adjusted risk in any age group in the Omicron wave compared to the Delta wave. Results for the outcome admission to hospital ≤14 days after positive test regardless of main cause were largely consistent with those for acute COVID-19 as main cause of hospitalisation, although we did observe a decreased risk in cases <18 years in the Omicron wave compared to Delta wave (aRR: 0.67, 95% CI: 0.48–0.94) (Table 2).

**Table 2.**
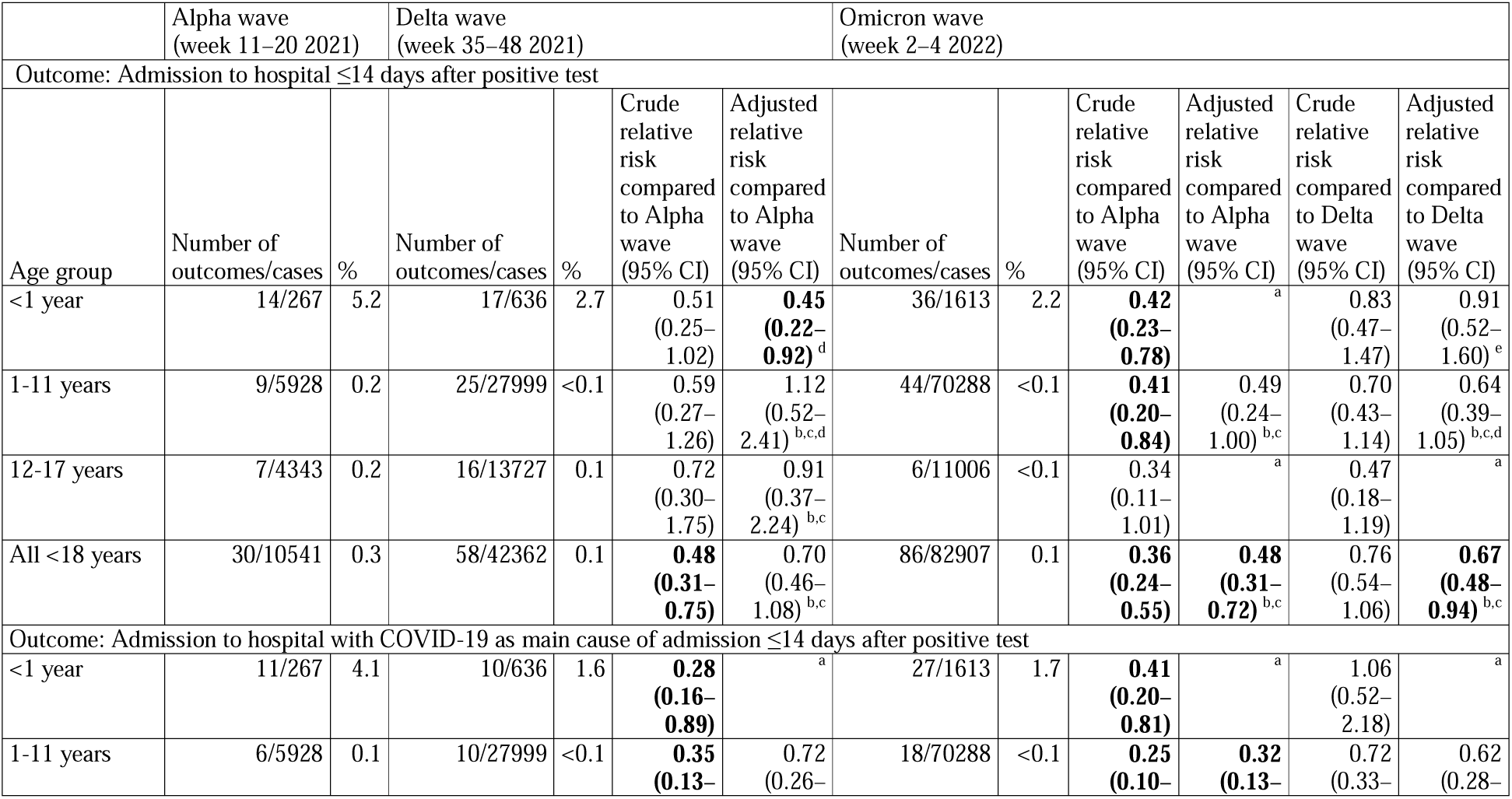
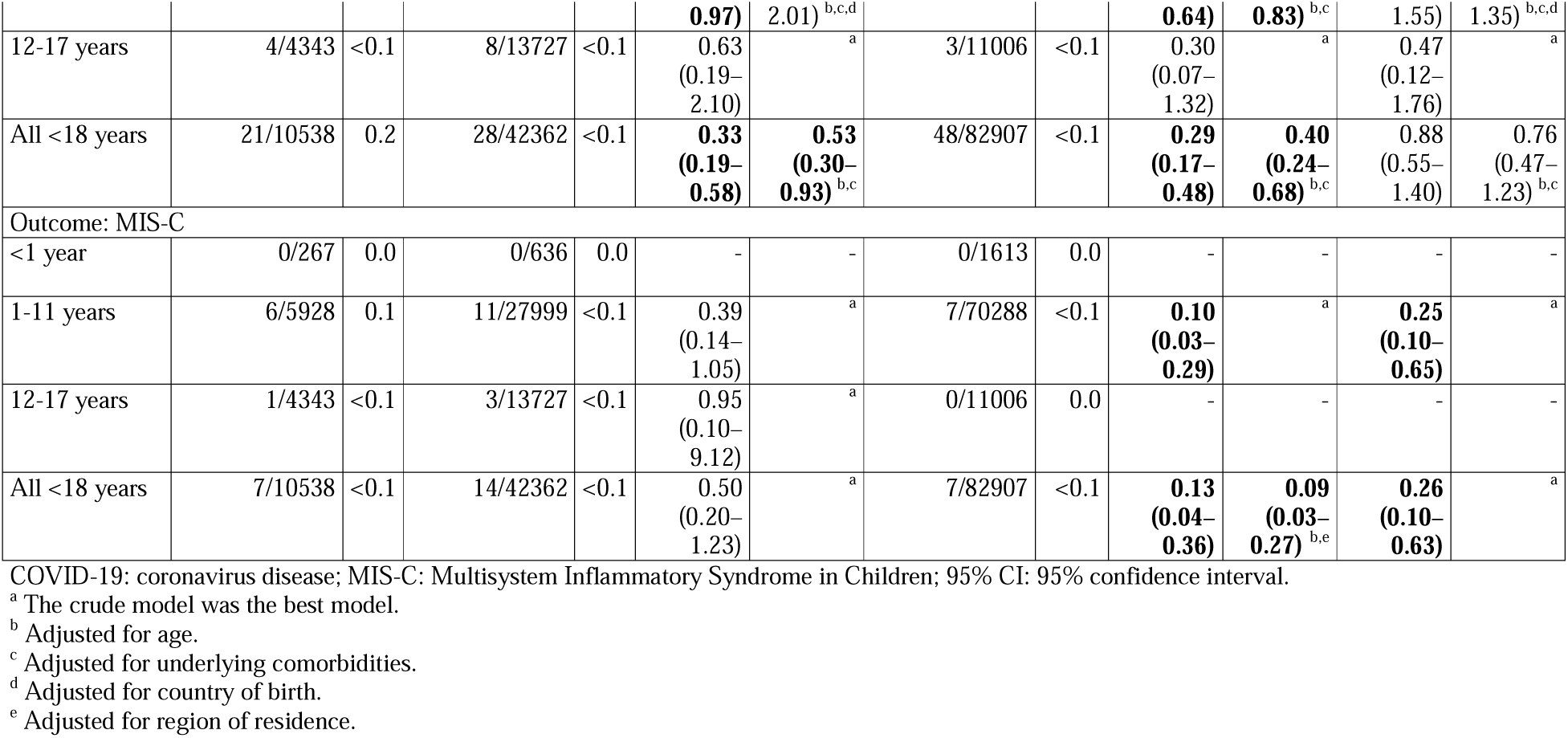
Number of reported cases admitted to hospital for acute COVID-19 or MIS-C, and crude and adjusted risk ratios from log-binomial regression, by age group and variant wave, cases <18 years who were unvaccinated and had also not previously been diagnosed with COVID-19, Norway, 15 March 2021 – 30 January 2022

### Risk of MIS-C

Twenty-eight cases of MIS-C were reported across the three waves (0.02% of all cases) (Table 2). The median age was 6.5 years (interquartile range (IQR): 4–9.5; range 1–17), 12 (43%) were female and 26 (93%) had no registered comorbidity. In the Alpha, Delta and Omicron waves, 7 (0.07%), 14 (0.03%) and 7 (0.008%) cases <18 years were diagnosed with MIS-C, respectively (Table 1, Table 2). We did not observe a significant difference in the risk of MIS-C in the Delta wave compared to the Alpha wave. The risk of MIS-C among cases <18 years was lower in the Omicron wave compared to the Alpha (aRR: 0.09, 95% CI: 0.03– 0.27) and Delta wave (aRR: 0.26, 95% CI: 0.10–0.63) (Table 2).

### Length of hospital stay, admission to intensive care and death

The median LoS among cases hospitalised with acute COVID-19 as main cause was 0.8 days (IQR: 0.4–3.2) in the Alpha wave, 1.4 days (IQR: 0.7–3.1) in the Delta wave and 1.1 days (IQR: 0.8–2.7) in the Omicron wave. Among the 28 MIS-C cases the median LoS was 4 days (interquartile range (IQR): 2–6). Data on LoS is not presented by wave for MIS-C cases due to the small number of MIS-C cases in each wave. Across all three waves, 6 (6%) cases hospitalised with acute COVID-19 as main cause and 4 (14%) MIS-C cases were admitted to ICU. This equates to 0.007% of reported cases being admitted to ICU for either acute COVID-19 or MIS-C in Norway. At the end of follow-up, there were no reported deaths among those hospitalised (either for acute COVID-19 or MIS-C), nor within 30 days of positive test among those not hospitalised.

## Discussion

We find no clear evidence of a difference in the risk of hospitalisation due to acute COVID-19 among cases <18 years in the Omicron wave compared to the Delta wave. Our results are in line with similar national studies from both England (5) and Denmark (12). The study in England, which included over 300,000 COVID-19 cases and 1,500 hospitalisations <20 years, did suggest a potentially small decrease in the risk of hospitalisation for Omicron compared to Delta ≤14 days after positive test among unvaccinated 10–19-year-olds (adjusted hazard ratio: 0.78, 95% CI: 0.60–1.00). We did find a lower risk for hospitalisation regardless of main cause in the Omicron wave compared to the Delta wave. However, our results for this indicator should be interpreted with caution as it will include patients admitted for non-COVID-19 related causes. We find no difference in the risk of hospitalisation with COVID-19 as main cause in the Omicron wave compared to the Delta wave.

The risk of hospitalisation with acute COVID-19 among cases <1 year was lower in the Delta and Omicron waves, compared to the Alpha wave. As the majority of hospitalised cases <1 year across the three waves were <3 months old, the decrease in risk may be related to protection offered to infants born to mothers vaccinated during pregnancy (27). Vaccination during pregnancy was first generally recommended in Norway before the start of the Delta wave. Conversely, we did not a see a difference in the risk of hospitalisation with acute COVID-19 among cases <1 year between the Omicron and Delta waves. This may reflect higher vaccination coverage among pregnant women, but lower vaccine effectiveness in the Omicron wave (7).

Compared to other VOC, the Omicron variant has been shown to replicate more easily in the upper respiratory tract but less well in the lungs (28). In this study we could not explore whether the clinical presentation of children hospitalised with Omicron had changed, compared to previous variant waves. There have been reports of an increase in upper respiratory complications, such as croup, among young children since Omicron became dominant (13, 14). Such changes in the clinical presentation of hospitalised paediatric COVID-19 patients would have important implications for future hospital capacity planning and management of paediatric patients as Omicron circulates.

Previous studies from the USA and Denmark estimated the incidence of MIS-C (defined based on the case definition from the Centers for Disease Control and Prevention) to be between 1 in 3,000 to 1 in 4,000 children infected with the wild-type SARS-CoV-2 variant (29, 30). A subsequent study from Denmark found that this risk did not change among unvaccinated cases during a wave of Delta infections (20). We find that the incidence of MIS-C (based on the case definition from the World Health Organisation) in Norway during the Delta wave was approximately 1 in 3,000 cases who were unvaccinated and had not been previously diagnosed with COVID-19. This decreased to 1 in 12,000 in the Omicron wave, an estimated 75% decrease (95% CI: 37%–90%) in risk. This may suggest a lower intrinsic risk of MIS-C for Omicron compared to Delta. This is an encouraging finding, especially given evidence that this risk may be further reduced through vaccination. Studies from Denmark and the USA have estimated the effectiveness of two doses of Comirnaty^®^ against MIS-C to be 91%–94% during periods of Delta dominance (20, 31). In this study, when we included vaccinated cases 12–17 years in a sensitivity analysis, no reported MIS-C cases had completed a two-dose primary vaccination series ≥7 days before positive test (supplement A, section 3). This is in line with another study from France (32). However, it remains to be seen if the same level of vaccine effect against MIS-C is maintained during a period of Omicron dominance. Omicron has been linked to an increase in breakthrough infections (7, 33) and lower vaccine effectiveness against some severity outcomes among children and adolescents 5–17 years (34). The proportion of MIS-C cases admitted to ICU in our study is notably lower than reported by others (11, 20, 35). Here, differences in the settings, definition of MIS-C cases and the definition of ICU admission need to be considered. For example, our definition of ICU admission will exclude stays in intermediate observation posts in a paediatrics unit.

In this study we have analysed national registry data from a setting with a broad COVID-19 testing strategy among children and adolescents. Hospitals in Norway functioned within capacity during the study period. Criteria for hospitalisation and isolation for COVID-19 patients, and clinical criteria for the diagnosis of MIS-C, were consistent.

An important limitation with severity studies based on reported COVID-19 cases is that undiagnosed cases will affect reported outcome proportions, while systematic differences in undiagnosed cases between groups may affect comparisons. In our study, this is particularly relevant for the comparison of the Alpha wave to other variant waves. The testing strategy was enhanced after the Alpha wave, thus a higher proportion of asymptomatic and mild cases may have been diagnosed among school-age children and adolescents in the Delta and Omicron waves. Also, the small number of outcomes must be considered, which restricted further exploration of our results. Given the low incidence of severe outcomes among child and adolescent COVID-19 cases, which may be further reduced through vaccination (20, 31, 34, 36), analyses of pooled data from several countries or meta-analyses could provide more precise estimates that can better elucidate differences in the risk of these outcomes between VOC in younger age groups. Further, we have based the analysis on variant waves, not cases with known data. However, in models for all COVID-19 cases reported in Norway, results based on these waves were consistent with analyses based on cases with known variant in periods when one variant was superseding another (see supplement A, section 4). Finally, we analysed an Omicron wave when the sublineage BA.1 was the dominant circulating variant. Further studies are still needed to establish differences in disease severity between different Omicron sublineages.

## Conclusion

We found no evidence of a difference in the risk of hospitalisation due to acute COVID-19 among unvaccinated cases <18 years for Omicron compared to Delta in Norway, but a reduced risk of hospitalisation among cases <1 year in Omicron and Delta waves, compared to the Alpha wave. Results also suggest a decrease in the risk of MIS-C among those infected with the Omicron variant compared to the Delta and Alpha variant.

## Data Availability

The dataset analyzed in the study contains individual-level linked data from various central health registries, national clinical registries, and other national administrative registries in Norway. The researchers had access to the data through the national emergency preparedness registry for COVID-19 (Beredt C19), housed at the Norwegian Institute of Public Health (NIPH). In Beredt C19, only fully anonymized data (i.e. data that are neither directly nor potentially indirectly identifiable) are permitted to be shared publicly. Legal restrictions therefore prevent the researchers from publicly sharing the dataset used in the study that would enable others to replicate the study findings. However, external researchers are freely able to request access to linked data from the same registries from outside the structure of Beredt C19, as per normal procedure for conducting health research on registry data in Norway. Further information on Beredt C19, including contact information for the Beredt C19 project manager and information on access to data from each individual data source, is available at https://www.fhi.no/en/id/infectious-diseases/coronavirus/emergency-preparedness-register-for-covid-19/.

## Abbreviations

VOC: Variant of concern
SARS-CoV-2: Severe acute respiratory syndrome coronavirus 2
COVID-19: Coronavirus disease
MIS-C: Multisystem inflammatory syndrome in children
LoS: Length of stay
ICU: intensive care unit
aRR: Adjusted risk ratio
CI: Confidence interval
PCR: Polymerase chain reaction

## Acknowledgements

First and foremost, we wish to thank all those who have helped establish and/or report data housed in the national emergency preparedness registry for COVID-19 at the Norwegian Institute of Public Health. We would also like to specifically thank Trude Lyngstad, Jostein Starrfelt, Elina Seppälä, Mari Grøsland and ‘Team Riskgroup’ at the Norwegian Institute of Public Health for their assistance in the cleaning of the data from the different registries, and additionally Trude Lyngstad for her assistance in the production of Figure S1.

## Notes

Conflict of interest: The authors declare that they have no competing interests.

### Competing Interest Statement

The authors have declared no competing interest.

### Funding Statement

This research was undertaken as part of routine work at the Norwegian Institute of Public Health. No specific funding was received.

