## Supplement A for "Severe outcomes in unvaccinated COVID-19 cases <18 years during different variant waves in Norway"

**1. National COVID-19 vaccine recommendations and vaccination coverage among persons 5–17 years in Norway**

In Norway, vaccination was first offered to children and adolescents with severe comorbidities; from February 2021 for 16–17-year-olds, June 2021 for 12–15-year-olds, and late December for 5–11-year-olds. Since August 2021, all 16–17-year-olds have been recommended vaccination. All 12–15-year-olds and 5–11-year-olds have been offered vaccination since September 2021 and January 2022, respectively (Table S1). As of March 2022, the mRNA vaccine Comirnaty® (BioNTech-Pfizer, Mainz, Germany/New York, United States) was the vaccine recommended to be administered to children <18 years as part of the national vaccination programme.

Among 16–17-year-olds, one-dose coverage increased from 1.5% in late-August to over 80% by late-October, reaching 89% by 31 January 2022. Two-dose coverage was <5% until late-October, before increasing to over 70% in late-December and reaching 79% by 31 January 2022 (Figure S1). Among 12–15-year-olds, one-dose coverage increased from <1.0% in early September to over 70% in early-December, reaching 75% by 31 January 2022. Two-dose coverage reached 5.4% by 31 January 2022 (Figure S1). At the end of January 2022, 1.0% of the national population aged 5–11 years had received one vaccine dose.

**Table S1. COVID-19 vaccine recommendations for children and adolescents 5–17 years up to January 2022, Norway**

| Age group | Recommendation for those with severe underlying disease | Recommendation/offer for all within age group |
| --- | --- | --- |
| 16–17 years | February 2021 | August 2021: 2 doses recommended, 8–12 weeks interval |
| 12–15 years | June 2021 | September 2021: 1st dose offered  January 2022: 2nd dose offered |
| 5–11 years | December 2021 | January 2022: 2 doses offered, 8–12 weeks interval |

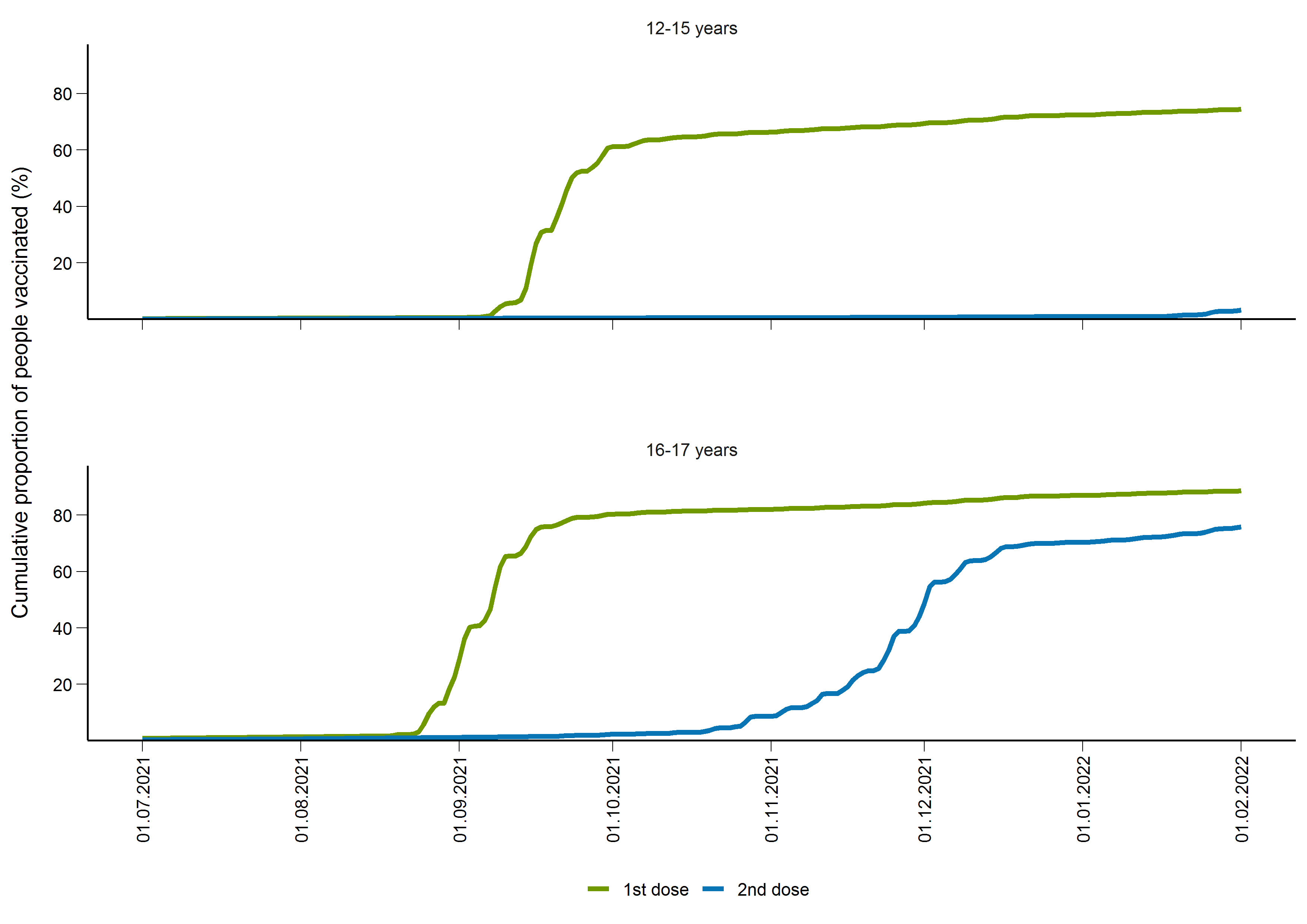

**Figure S1. National vaccination coverage of COVID-19 vaccines in Norway by age group, number of doses and date, persons 12–17 years, 1 July 2021 – 31 January 2022.**

**2. Additional information on the data sources and definitions**

All data in this study came from the [national emergency preparedness register](https://www.fhi.no/en/id/infectious-diseases/coronavirus/emergency-preparedness-register-for-covid-19/), Beredt C19. Beredt C19 contains individual-level data from central health registries, national clinical registries, and other national administrative registries.

*1.1 Reported cases of COVID-19*

We included data on reported cases of laboratory-confirmed SARS-CoV-2 infection from the Norwegian Surveillance System for Communicable Diseases (MSIS). Up to 24 January 2022, reinfections were registered if there were ≥6 months between two positive sampling dates for an individual. This will thus exclude reinfections within a 6-month period, of which the Omicron variant could be of higher risk (1, 2). From 25 January 2022, reinfections were registered if there were ≥2 months between two positive sampling dates for an individual.

*1.2 Laboratory testing for variants*

Data on virus variants came from the MSIS laboratory database (national laboratory database), which receives SARS-CoV-2 test results from all Norwegian microbiology laboratories. Variants are identified based on whole genome sequencing, Sanger partial S-gene sequencing or PCR screening targeting specific single nucleotide polymorphisms, insertions or deletions. The laboratory testing for variants of SARS-CoV-2 in Norway has been described in further detail elsewhere (3).

*1.3 National identity number*

Data on persons with a national identity number was drawn from the national population registry (Folkeregistret). The national identity number was essential to link data from all registries used in the analysis. The national population registry was also used to identify all deaths in our study cohort during the study period.

*1.4 Hospitalisation and intensive care admission with acute COVID-19*

We obtained data on hospitalisation following a positive PCR test for SARS-CoV-2 from the Norwegian Intensive Care and Pandemic Registry (NIPaR). All Norwegian hospitals report to NIPaR, and reporting is mandatory. Hospitals in Norway functioned within capacity during each variant wave in the study period, and criteria for hospitalisation and isolation for COVID-19 patients were consistent.

For patients who contracted COVID-19 while admitted to hospital, the time of admission is set to the date of symptom onset, or date of sampling if the patient is asymptomatic. The reported main cause of hospitalisation is a clinical assessment. For patients reported with a different main cause than COVID-19, we cannot rule out that COVID-19 may have been a contributing factor for admission. There is no reason to believe, however, that this assessment would differ between patients infected with different variants or hospitalised in different periods. Full details on the registration of hospitalised patients are available here (in Norwegian): <https://helse-bergen.no/norsk-pandemiregister/registrering-i-norsk-pandemiregister-informasjon-til-ansatte>

NIPaR also includes data on patients who have tested positive for COVID-19 and are admitted to an intensive care unit (ICU). Patients are registered as ICU patients if they fulfil one of five categories:

1. LoS over 24 hours in intensive care
2. Require mechanical ventilation
3. Are transferred between intensive care wards
4. Persistent administration of vasoactive medication
5. LoS under 24 hours, but passed away during stay in intensive care

Full details on the registration of ICU patients are available here (in Norwegian): <https://helse-bergen.no/norsk-pandemiregister/registrering-i-norsk-pandemiregister-informasjon-til-ansatte>.

*1.5 Cases of multisystem inflammatory syndrome in children*

The Norwegian Patient Register (NPR) contains health information about all persons who have received treatment, or who are waiting for treatment in public specialist health-care services, including private institutions and medical specialists contracted to the regional health authorities. Admission to hospital with multisystem inflammatory syndrome in children (MIS-C) was defined as patients registered in NPR with the ICD-10 code U10.9. Clinical criteria for the diagnosis of MIS-C cases in Norway are based on World Health Organisation guidelines (4).

*1.6 Vaccination status*

Data on COVID-19 vaccinations came from the Norwegian Immunisation Registry, SYSVAK. Unvaccinated cases were those who were unvaccinated with a COVID-19 vaccine at date of positive test, and who had also not been previously diagnosed with COVID-19, as reported to MSIS.

*1.7 Underlying comorbidities*

Data on underlying comorbidities, as stipulated by the [national COVID-19 vaccination programme](https://www.fhi.no/en/id/vaccines/coronavirus-immunisation-programme/who-will-get-coronavirus-vaccine-first/), was based on ICD-10 codes from NPR, and ICPC-2 codes from the Norway Control and Payment of Health Reimbursement database (Table S1). Underlying comorbidities that have been defined as increasing the risk of severe COVID-19 were divided into two groups.

- Medium-risk includes people with diseases/conditions that entail a moderate risk of severe COVID-19. This includes chronic liver disease or significant hepatic impairment, immunosuppressive therapy as in autoimmune diseases, diabetes, chronic lung disease including cystic fibrosis and severe asthma which have required the use of high dose inhaled or oral steroids within the past year, obesity with a body mass index (BMI) of ≥35 kg/m2, dementia, chronic heart and vascular disease (with the exception of high blood pressure) and stroke.
- High-risk includes people with diseases/conditions that carry a high risk of severe COVID-19, also in younger individuals. These comorbidities include having received an organ transplant, immunodeficiency, hematological cancer in the last five years, other active cancers, ongoing or recently discontinued treatment for cancer (especially immunosuppressive therapy, radiation therapy to the lungs or cytotoxic drugs), neurological or neuromuscular diseases that cause impaired cough or lung function (e.g., ALS and cerebral palsy), Down syndrome and chronic kidney disease, or significant renal impairment.

**Table S1. ICD-10 codes from the Norwegian Patient Registry and ICPC-2 codes from the Norway Control and Payment of Health Reimbursement database used to identify cases with underlying comorbidities**

| Underlying comorbidity | Specifications | ICD10-codes | ICPC-2 codes |
| --- | --- | --- | --- |
| Cardiovascular diseases, not including hypertension |  | I05, I06, I07, I08, I09, I2, I31,  I32, I34, I35, I36, I37, I39, I40, I41, I42, I43, I46, I48, I49, I50, I60, I61, I62, I63, I64, I69.1, I69.2, I69.3, I69.4, I69.8, I69.0 | K74, K75, K76, K77, K78, K82, K83, K90, K91 |
| Chronic pulmonary diseases, including asthma |  | J41, J42, J43, J44, J45, J46,  J47, J84, J98, E84 | R95, R96 |
| Compromised immune function | Organ transplantation, immune deficiency disorders, autoimmune conditions treated with immunosuppressants | Z94.0, Z94.1, Z94.2, Z94.3, Z94.4, Z94.8, D80, D81, D82, D83, D84, G35, M05, M08, M06, M07, M09, M13, M14, K50, K51 |  |
| Neurological and musculoskeletal disorders with compromised lung or cough function |  | G1, G20, G21, G23, G24, G40.5, G61.0, 70, G71, G80.0, G80.2, G80.3, F72, F73, F84.0, F84.1, Q05.0, Q05.1, Q05.2, Q05.3, Q05.04, Q05.5, Q05.6 |  |
| Diabetes |  | E10, E11, E12, E13, E14 | T89, T90 |
| Active cancer treatment or hematological cancer |  | C81, C82, C83, C84, C85, C86, C87, C88, C89, C90, C91, C92, C93, C94, C95, C96, D45, D45, D47, C0, C1, C2, C3, C4, C5, C6, C7, C80, D32, D33, D35.2, D35.3, D35.4, D42, D43, D44.2, D44.3, D44.4 |  |
| Other risk groups | Dementia, chronic kidney and liver disease, obesity | N18.3, N18.4, N18.5, K70.4,  K72, F00, F01, F02, F03, G30, G31, E66 | P70, T82 |

**3. Estimates among all reported COVID-19 cases 12–17 years, compared to estimates among unvaccinated cases, Omicron wave compared to Delta wave**

**Table S2. Number of reported cases admitted to hospital for acute COVID-19 or MIS-C, and crude and adjusted risk ratios from log-binomial regression, by age group, vaccination status and variant wave, cases 12–17 years, Norway**

|  | Delta wave  (week 35–48 2021) | | Omicron wave  (week 2–4 2022) | | | |
| --- | --- | --- | --- | --- | --- | --- |
| Age group | Number of outcomes/cases | % | Number of outcomes/cases | % | Crude relative risk compared to Delta wave (95% CI) | Adjusted relative risk compared to Delta wave (95% CI) |
| Outcome: Admission to hospital ≤14 days after positive test | | | | | | |
| Unvaccinated only (main analysis) | 16/13727 | 0.1 | 6/11006 | <0.1 | 0.47  (0.18–1.19) | ^a^ |
| All cases, including vaccinated and previous infections | 20/24888 | <0.1 | 27/58756 | <0.1 | 0.57  (0.32–1.02) | 0.83  (0.42–1.64) ^b,c^ |
| Outcome: Admission to hospital with COVID-19 as main cause of admission ≤14 days after positive test | | | | | | |
| Unvaccinated only (main analysis) | 8/13727 | <0.1 | 3/11006 | <0.1 | 0.47  (0.12–1.76) | ^a^ |
| All cases, including vaccinated and previous infections | 10/24888 | <0.1 | 7/58756 | <0.1 | **0.30**  **(0.11–0.78)** | 0.45  (0.15–1.36) ^c^ |
| Outcome: MIS-C | | | | | | |
| Unvaccinated only (main analysis) | 3/13727 | <0.1 | 0/11006 | 0.0 | - | - |
| All cases, including vaccinated and previous infections ^d^ | 5/24888 | <0.1 | 2/58756 | <0.1 | **0.16**  **(0.03–0.87)** | ^a^ |

COVID-19: coronavirus disease; MIS-C: Multisystem Inflammatory Syndrome in Children; 95% CI: 95% confidence interval.

^a^ The crude model was the best model.

^b^ Adjusted for age.

^c^ Adjusted for vaccination status (vaccinated with at least one dose ≥21 days before positive test or previously diagnosed with COVID-19 vs. other).

^d^ Among the four additional MIS-C cases in the ‘All cases’ cohort, none had completed a two-dose primary vaccination series ≥7 days before positive test.

This sensitivity analysis was not run for the age group 1–11 years, as only 92 cases total in this age group (all but one in the Omicron wave) had received at least one vaccine dose before positive test.

**4. Estimates among all reported COVID-19 cases by variant wave, compared to cases with known variant**

In order to validate the variant wave variable, we ran models for all COVID-19 cases reported in Norway and compared these to analyses based on cases with known variant during periods when one variant was in the process of superseding another (Table S4 and Table S5). We could not run models based on cases with known variant for just those 0–17 years, due to the small number of cases with known variant in this age group. In the analysis of cases with known variant, the alpha–delta period was week 18–32 2021 and the delta–omicron period week 49 2021–week 1 2022, as used in previous analyses of cases with known variant (5, 6). The variant wave variables were defined as in the main manuscript. We ran a logistic regression model and estimated odds ratios, as opposed to log-binomial regression or cox regression. Cox regression was not possible as the variant wave variable violated the proportional hazards assumption, while a log-binomial model did not converge in the variant wave analysis. The models are adjusted for variant (wave or known), age group (<1, 1-5, 6-11, 12-15, 16-17, 18-29, 30-44, 45-54, 55-64, 65-74, 75+), gender, vaccination status, county of residence, underlying comorbidities and country of birth. Vaccination status, county of residence, underlying comorbidities and country of birth were defined as in (6). The models for known variant are also adjusted for sampling week, which was not included in the variant wave models due to collinearity. We analysed three different outcomes to see if our results remained robust. Results were consistent, regardless of whether variant wave or known variant was analysed. Results were also consistent with our previous comparison of delta and alpha cases using log-binomial regression (adjusted relative risk: 0.97, 95% CI: 0.76–1.23) (5) and delta and omicron cases using cox regression (adjusted hazard ratio: 0.27; 95% CI: 0.20–0.36) (6), where the outcome was admission with COVID-19 as main cause of admission.

**Table S4. Crude and adjusted odds ratios from logistic regression for different outcomes among COVID-19 cases, by variant wave or cases with known variant, Delta variant compared to Alpha variant, Norway**

| Outcome | Analysis of variant data | Alpha | | Delta | | | |
| --- | --- | --- | --- | --- | --- | --- | --- |
|  |  | Number of outcomes/cases | % | Number of outcomes/cases | % | Crude odds risk compared to Alpha wave (95% CI) | Adjusted odds risk compared to Alpha wave (95% CI) |
| Admission to hospital within 14 days of positive test | Variant wave | 1440/39523 | 3.7 | 2074/127358 | 1.6 | **0.44**  **(0.41–0.47)** | 1.06  (0.97–1.16) |
|  | Cases with known variant | 281/12162 | 2.3 | 133/8227 | 1.6 | **0.69**  **(0.56–0.86)** | 0.77  (0.51–1.18) |
| Admission to hospital with COVID-19 as main cause of admission within 14 days of positive test | Variant wave | 1218/39523 | 3.1 | 1505/127358 | 1.2 | **0.38**  **(0.35–0.41)** | 0.96  (0.87–1.06) |
|  | Cases with known variant | 242/12162 | 2.0 | 108/8227 | 1.3 | **0.66**  **(0.52–0.82)** | 0.80  (0.51–1.28) |
| Admission to hospital with COVID-19 as main cause of admission or death within 28 days of positive test | Variant wave | 1283/39523 | 3.3 | 1770/127358 | 1.4 | **0.42**  **(0.39–0.45)** | 1.00  (0.90–1.10) |
|  | Cases with known variant | 256/12162 | 2.1 | 120/8227 | 1.5 | **0.69**  **(0.55–0.86)** | 0.75  (0.48–1.18) |

95% CI: 95% confidence interval. Bold = statistically significant.

**Table S5. Crude and adjusted odds ratios from logistic regression for different outcomes among COVID-19 cases, by variant wave or cases with known variant, Omicron variant compared to Delta variant, Norway**

| Outcome | Analysis of variant data | Delta | | Omicron | | | |
| --- | --- | --- | --- | --- | --- | --- | --- |
|  |  | Number of outcomes/cases | % | Number of outcomes/cases | % | Crude odds risk compared to Delta wave (95% CI) | Adjusted odds risk compared to Delta wave (95% CI) |
| Admission to hospital within 14 days of positive test | Variant wave | 2074/127358 | 1.6 | 951/321874 | 0.3 | **0.18**  **(0.17–0.19)** | **0.43**  **(0.39–0.47)** |
|  | Cases with known variant | 756/51995 | 1.5 | 194/40641 | 0.5 | **0.33**  **(0.28–0.38)** | **0.33**  **(0.27–0.41)** |
| Admission to hospital with COVID-19 as main cause of admission within 14 days of positive test | Variant wave | 1505/127358 | 1.2 | 457/321874 | 0.1 | **0.12**  **(0.11–0.13)** | **0.29**  **(0.26–0.33)** |
|  | Cases with known variant | 551/51995 | 1.1 | 107/40641 | 0.3 | **0.25**  **(0.20–0.30)** | **0.27**  **(0.21–0.36)** |
| Admission to hospital with COVID-19 as main cause of admission or death within 28 days of positive test | Variant wave | 1770/127358 | 1.4 | 559/321874 | 0.2 | **0.12**  **(0.11–0.14)** | **0.31**  **(0.28–0.35)** |
|  | Cases with known variant | 611/51995 | 1.2 | 126/40641 | 0.3 | **0.26**  **(0.22–0.32)** | **0.27**  **(0.21–0.35)** |

95% CI: 95% confidence interval. Bold = statistically significant.
